# Projection of hospitalization by COVID-19 in Brazil following different social distance policies

**DOI:** 10.1101/2020.04.26.20080143

**Authors:** Henrique Matheus Ferreira da Silva, Rafael Silva Pereira, Raquel de Abreu Junqueira Gritz, Adolfo Simões, Fabio Porto

## Abstract

Following the first infections in the Wuhan City, COVID-19 became a global pandemic as declared by the World Health Organization (WHO). Since it is an airborne disease transmitted between humans, many countries adopted a quarantine on their own population as well as closing borders measures. In Brazil many are discussing the best way to manage the opening of the quarantine under the constraints of hospitals infrastructure. In this work we implement a forecast of the demand for hospital beds for the next 30 days for every state in Brazil and under different quarantine flexibilization scenarios and analyse how long it would take for the demand to exceed current available hospital beds.

## 1. INTRODUCTION

COVID-19, a disease caused by the SARS-CoV-2 virus, was first identified on December 31, 2019 in Wuhan City, and within weeks, it became a disease recognized as a global pandemic [14] by the World Health Organization. The first confirmed infection in Brazil occurred in São Paulo, on February 26, 2020, and later spread throughout the country. Subsequently its arrival in Brazil, the quarantine of the population was determine due to the high rate of virus transmission and lack of a vaccine, which could result in high overcrowding of the health system. After more than a month of the adoption of the quarantine in Brazil, dialogues are occurring about its possible future.

Given this scenario, this article consists in a prediction for the next 30 days of the advancement of COVID-19 in Brazil by state, predicting the point at which the demand of hospital bed for patients in critical condition would surplus the installed capacity at each state, based on 3 scenarios of social distance policies. Quarantine scenarios were based on the adoption of public policies on 3 countries [?] in their first 30 days of infection: South Korea (quarantine with more incisive policies, with the inexistence of prohibition on populational movement)[3]; Germany (quarantine with more balanced public policies, recommending the use alcohol gel, masks and avoiding agglomerations, but without prohibiting the free movement of the population)[2]; Italy (tenuous quarantine, without any countermeasure)[11],[9],[8]. For each one of these policies, we predict the number of patients requiring hospitalization per state, and confront then with the corresponding number of available beds, obtained from the official database CNES[7] [1]. We remove from these datasets the pediatric and neonatal beds, and assume that any other bed could receive a patient affected by the COVID-19. Given the number of predicted patients requiring hospitalization and the number of available beds, we identify whether the supply would fulfil the demand. The remaining of the paper is structured as follows. In session II, we discuss the methodology. Then, section III presents the results, and section IV concludes.

## 2. METHODOLOGY

In this work we propose to evaluate the demand for hospitalization due to the COVID-19 spread at each state in Brazil. To do so, we gathered data about COVID-19 spread over the world from the John Hopkins resource center [12]. Data about the spread at each Brazilian state was gathered from official sources [13]. The amount of available beds for COVID-19 patients was obtained from the CNES database on DATASUS [7] and [1]. After obtaining the data the following procedure is applied:

### 2.1 Statistical analysis

From the John Hopkins Coronavirus Resource Center data, we subset only the information regarding the evolution in number of confirmed infected patients, and the number of deaths, in the three countries considered in our scenarios: South Korea, Germany, and Italy. The number of confirmed and dead cases are then normalized by dividing them by the population size of the respective country. This way we can compare different countries since we will be looking at the percentage of the infected population.

Since COVID-19 has as its transmission vectors human themselves, it most basic model is an exponential function given by the solution of the equation 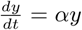, where *α* denotes the infection rate of the disease. This model of course does not account for recovery or deaths from the disease. It also does not consider other measures that can be done to suppress the infection spread. Once by taking into account these different scenarios the predictions would be affected. Based on this assumption, in order to circumvent this problem we calculate the infection rate increase as defined by equation: 1

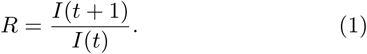

where *t* +1 denotes the next day. For each scenario, we calculate *R* above during the first 31 days and from this we calculate the cumulative product of *R*. This stores the percentage of the infected increase based on the number of infected already known. The resulting value does not take into account deaths or recovery, to take these into account the number of active people with the disease is given by formula 2:

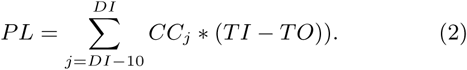

In which PL represents the projection of the number of beds in need, DI is the number of days to occur hospitalization(10 days), CC is the number of confirmed cases of COVID-19, TI is the hospitalization rate of people affected with COVID-19 and TO is the rate of death (3.2%) estimate given by the World Health Organization, (WHO)[4],[5],[10], at the time of this writing.

Since we are interested in analyzing how long would it take for the capacity of hospitals to be over passed, we assume that the number of patients in hospitals, who would need beds, to be 5% [6] of the active infected population, as denoted in 2. So we can predict, for each state, whether or when the demand for hospital beds exceeds the supply.

### 2.2 Obtaining hospital beds data

Observing the equation that predicts the increase in the number of people infected by COVID-19 needing hospitalization for each Brazilian state, and considering to be known the number of available hospital beds, it is possible to identify when the health facilities at each state would fall short on the demand, notwithstanding, offering the conditions to treat other types of diseases.

In this context, using the best-case scenario as a premise, we assume that all hospital beds are available for the hospitalization of patients infected by COVID-19. Thus, for each Brazilian state, Intensive Care Unit (ICU) beds, as well as general hospital beds, consisting of beds for clinical and surgical hospitalization, were considered in this study as available for hospitalization. We only eliminated from the set of available beds, the pediatric and neonatal. It is worth mentioning that, given the high demand for care and the number of beds available, it is assumed that all beds are considered for hospitalization and that they offer initial support for treatment in the event of a full ICU bed.

## 3. RESULTS

Assuming April 22, 2020 as the milestone of the adoption of the elected quarantine scenarios, we created the projection on the need for hospital beds for the following 30 days. Figures 1,2,3,4,5 present the states with the most alarming scenarios, in which analyzing the upper part of the confidence interval, even if all the beds available today could accommodate those infected by COVID-19, the health system would collapse in all three quarantine scenarios within this time window.

**Figure 1:**
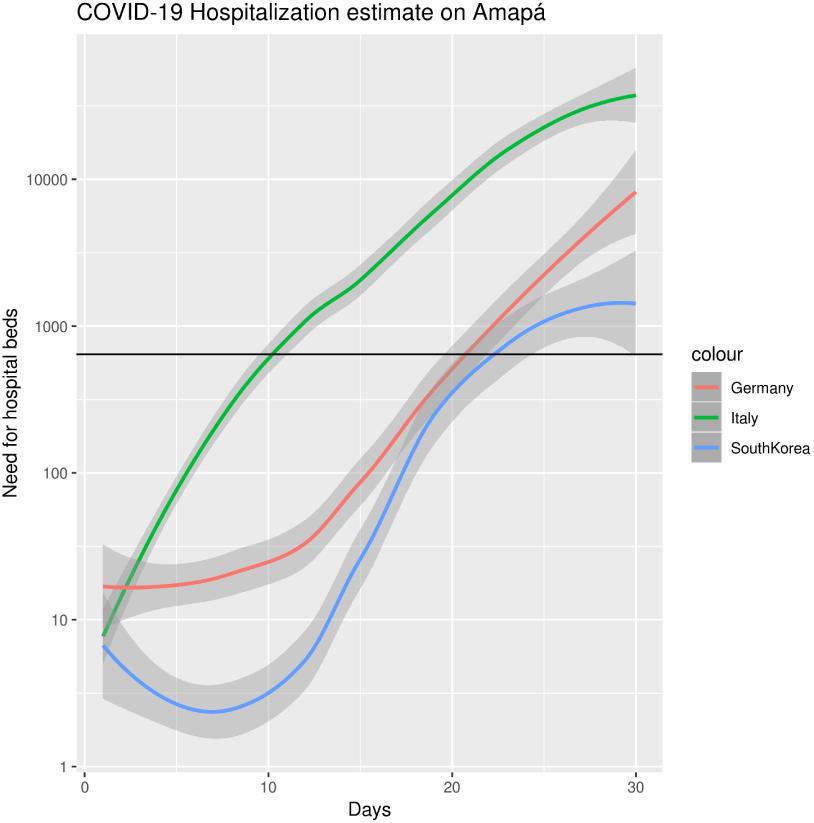
Projection of the need for hospitalization beds over time in the Amapá state

**Figure 2:**
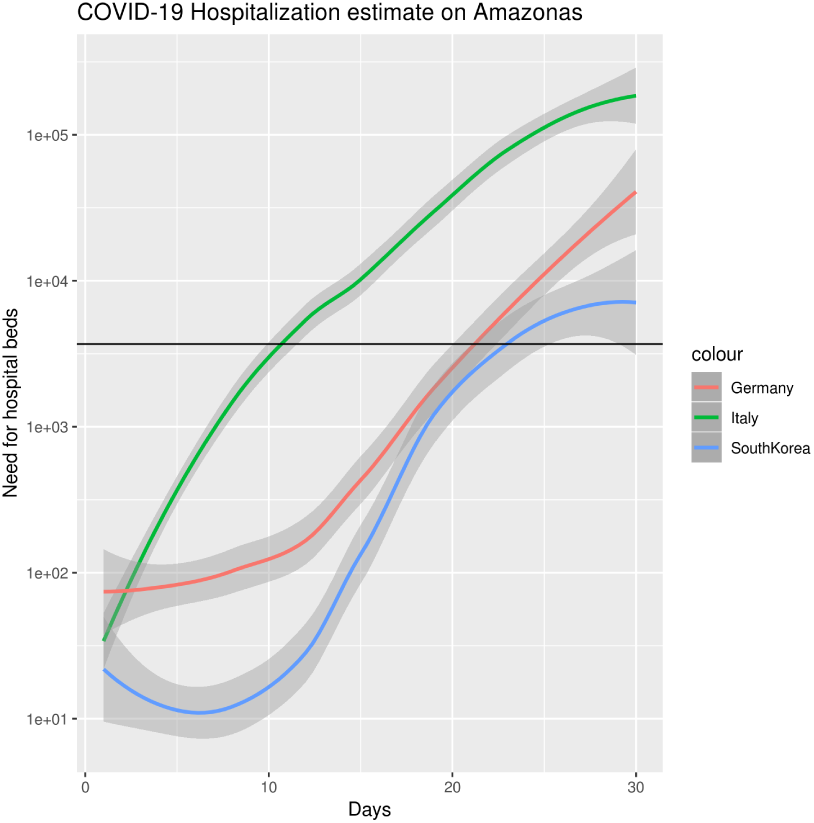
Projection of the need for hospitalization beds over time in the Amazonas state

**Figure 3:**
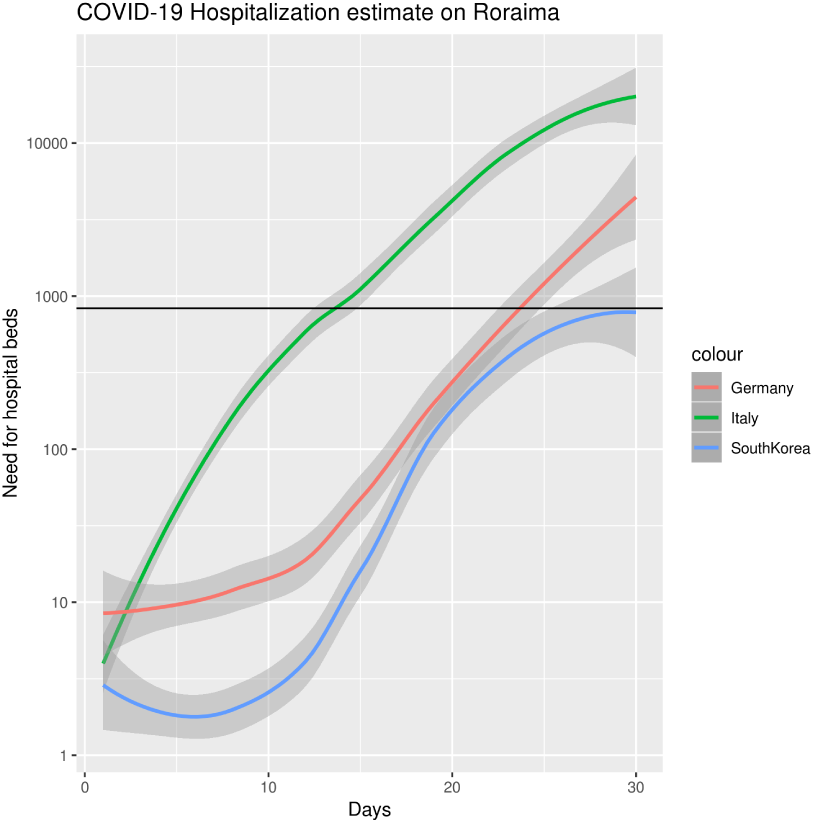
Projection of the need for hospitalization beds over time in the Roraima state

**Figure 4:**
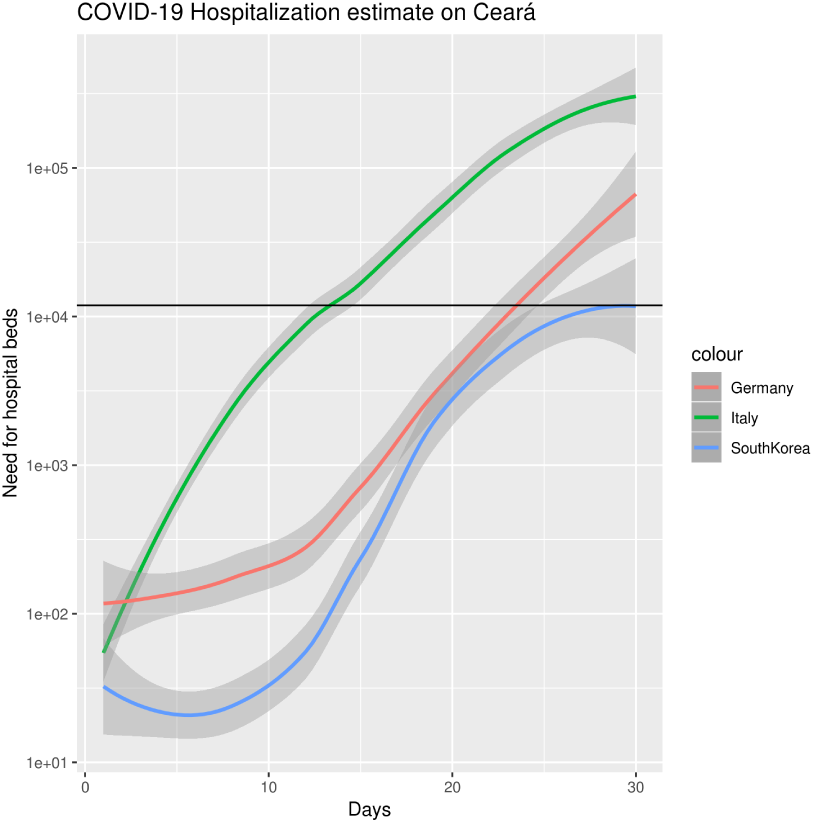
Projection of the need for hospitalization beds over time in the Ceará state

**Figure 5:**
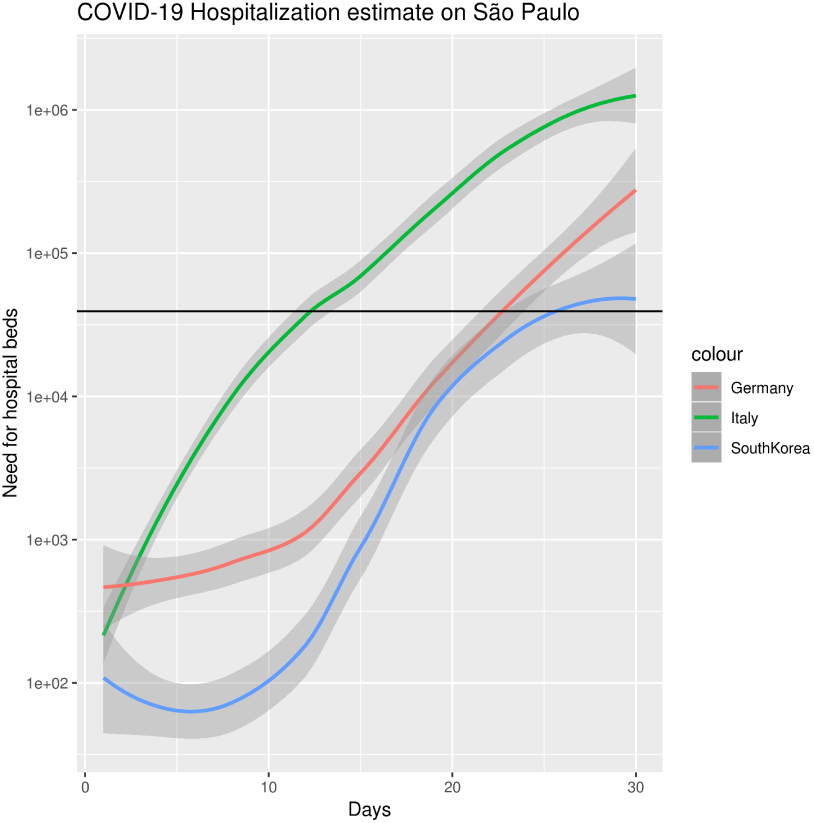
Projection of the need for hospitalization beds over time in the São Paulo state

Table 1 shows the point(in days) when states would collapse their health system, where values of - indicate that it would take more than 30 days for it to reach the state of overcrowding, analyzing the medium value in the confidence interval.

**Table 1:** Number of days from which the demand for hospitalization beds would surpass the existing installed capacity for COVID-19 patients for each studied quarantine scenario.

| Italy | Germany | SouthKorea | State |
| --- | --- | --- | --- |
| 17 | 26 | - | Distrito Federal |
| 22 | - | - | Goiás |
| 22 | - | - | Mato Grosso |
| 22 | - | - | Mato Grosso do Sul |
| 22 | - | - | Alagoas |
| 20 | 29 | - | Bahia |
| 14 | 26 | 29 | Ceará |
| 17 | 26 | - | Maranhão |
| 22 | - | - | Paraíba |
| 16 | 26 | - | Pernambuco |
| 22 | - | - | Piauí |
| 18 | 27 | - | Rio Grande do Norte |
| 22 | - | - | Sergipe |
| 16 | 26 | - | Acre |
| 11 | 21 | 23 | Amapá |
| 11 | 22 | 24 | Amazonas |
| 19 | 28 | - | Pará |
| 22 | - | - | Rondônia |
| 14 | 26 | - | Roraima |
| 22 | - | - | Tocantins |
| 16 | 26 | - | Espírito Santo |
| 22 | - | - | Minas Gerais |
| 16 | 26 | - | Rio de Janeiro |
| 13 | 24 | 26 | São Paulo |
| 22 | - | - | Paraná |
| 22 | - | - | Rio Grande do Sul |
| 19 | 29 | - | Santa Catarina |

## 4. CONCLUSION

By estimating the need for beds by Brazilian states, even adopting the best possible scenario with the population hospitalized for COVID-19 being discharged in a period of 10 days, in addition to the premise that all hospital beds (except for neonatal and pediatric) would be willing to receive the infected population, 5 states would collapse in the model adopted by South Korea, 16 states would collapse following the Germany scenario and all Brazilian states would collapse their health system based on the Italian scenario, in a 30 day time window. Therefore, it is recommended the adoption of public policies implemented by South Korea, in addition to increasing the number of beds with public policies with field hospitals (emergency beds for COVID-19).

It is noteworthy that in this scenario the Brazilian health system would make available for COVID-19 patients all available hospital beds, which would result in the said secondary deaths to COVID-19, people who would need a bed to be hospitalized due to other diseases (heart attack, car accident, stroke, etc.), but because of the overcrowd of the public health system would return home, resulting in an end that could have been contoured if hospital treatment had been given to the patient.

## Data Availability

We declare all links mentioned in the article available

https://www.reuters.com/article/us-china-health-italy/two-first-coronavirus-cases-confirmed-in-italy-prime-minister-idUSKBN1ZT31H

https://www.sciencemag.org/news/2020/03/coronavirus-cases-have-dropped-sharply-south-korea-whats-secret-its-success

https://www.worldometers.info/coronavirus/coronavirus-death-rate/

https://www.who.int/emergencies/diseases/novel-coronavirus-2019/situation-reports

https://www.who.int/dg/speeches/detail/who-director-general-s-opening-remarks-at-the-media-briefing-on-covid-19---3-march-2020

http://tabnet.datasus.gov.br/cgi/tabcgi.exe?cnes/cnv/leiintbr.def

http://tabnet.datasus.gov.br/cgi/tabcgi.exe?cnes/cnv/leiintbr.def

https://github.com/CSSEGISandData/COVID-19/tree/master/csse_covid_19_data/csse_covid_19_daily_reports

http://www.sciencedirect.com/science/article/pii/S1471491420300654

